# Maternal immune response and placental antibody transfer after COVID-19 vaccination across trimester and platforms

**DOI:** 10.1101/2021.11.12.21266273

**Authors:** Caroline G. Atyeo, Lydia L. Shook, Sara Brigida, Rose M. De Guzman, Stepan Demidkin, Cordelia Muir, Babatunde Akinwunmi, Arantxa Medina Baez, Erin McSweeney, Madeleine Burns, Ruhi Nayak, Maya K. Kumar, Chinmay D. Patel, Allison Fialkowski, Dana Cvrk, Ilona T. Goldfarb, Lael M. Yonker, Alessio Fasano, Michal A. Elovitz, Kathryn J. Gray, Galit Alter, Andrea G. Edlow

## Abstract

The availability of three COVID-19 vaccines in the United States provides an unprecedented opportunity to examine how vaccine platforms and timing of vaccination in pregnancy impact maternal and neonatal immunity. Here, we characterized the antibody profile after Ad26.COV2.S, mRNA-1273 or BNT162b2 vaccination in 158 pregnant individuals, and evaluated transplacental antibody transfer by profiling maternal and umbilical cord blood in 175 maternal-neonatal dyads. These analyses revealed lower vaccine-induced functions and Fc-receptor binding after Ad26.COV2.S compared to mRNA vaccination, and subtle advantages in titer and function with mRNA-1273 versus BN162b2. mRNA vaccinees had higher titers and functions against SARS-CoV-2 variants of concern. First and third trimester vaccination resulted in enhanced maternal immune responses relative to second trimester. Higher cord:maternal transfer ratios following first and second trimester vaccination reflect placental compensation for waning maternal titers. These results support vaccination early in pregnancy to maximize maternal protection throughout gestation, without compromising neonatal antibody protection.

## Introduction

Pregnant individuals with COVID-19 are not only at increased risk for severe morbidity and mortality,^1–4^ but also for adverse pregnancy outcomes including preterm delivery, pregnancy loss and stillbirth.^5–7^ While vaccination against COVID-19 is a critically important public health strategy to protect pregnant individuals and their pregnancies, only 33% of pregnant individuals have been vaccinated to date.^8^ Because pregnant individuals were excluded from initial vaccine clinical trials,^9–11^ data to guide clinical decision-making in this population have lagged behind those for the general population, contributing to vaccine hesitancy. One critical way to increase vaccine confidence in pregnant individuals is through observational data collected from individuals who have received the COVID-19 vaccine during pregnancy. To date, studies have demonstrated that pregnant people mount robust immunological responses to COVID-19 mRNA vaccines (BNT162b2 and mRNA-1273) with final titers achieved being comparable to those in non-pregnant women of reproductive age,^12–14^ and with similar safety and reactogenicity profiles.^12,14,15^ Several studies of pregnant people receiving COVID-19 mRNA vaccines primarily in the third trimester have also demonstrated the presence of anti-SARS-CoV-2-specific antibodies capable of neutralization and immune effector functions in umbilical cord blood at delivery,^12–14,16–19^ Maternal vaccination against COVID-19 has the potential not only to protect the pregnant woman, but to confer fetal and neonatal benefit by preventing adverse pregnancy outcomes related to severe maternal COVID-19 illness, and by providing newborns with immunity through transplacental and breastmilk transfer of maternal antibodies.^20,21^

Little is known, however, regarding how trimester-specific pregnancy immunity and different COVID-19 vaccine platforms may interact to impact maternal and neonatal protection from COVID-19. Trimester-specific immunological adaptations occur during normal pregnancy to promote implantation, support fetal growth and development, and stimulate parturition.^22–25^

Although COVID-19 vaccine safety across all trimesters of pregnancy has been well demonstrated,^15,26–28^ whether trimester of vaccination impacts vaccine immunogenicity or transplacental transfer to the neonate remains incompletely understood, as many pregnancies in which vaccination occurred in the first and early second trimester were ongoing at the time of initial study publications.^12–14,16–18^ Furthermore, data evaluating immune responses across COVID-19 vaccine platforms are limited even in the non-pregnant population,^29–32^ and platform-specific immune responses in the pregnant population have been limited to one comparison of mRNA-1273 vs BNT162b2.^13^ To date, no studies have directly assessed the immunogenicity of the Ad26.CoV.2 vaccine in pregnancy, nor compared these profiles to mRNA vaccines across gestation. To address these gaps, we used an unbiased systems serology approach to characterize the maternal antibody response and transplacental antibody transfer to umbilical cord by vaccine platform (BNT162b2, mRNA-1273, or Ad26.CoV2) and by trimester of vaccination.

## Results

### Clinical and demographic information

Using systems serology, we profiled the vaccine-induced immune response in a cohort of 158 women who completed a COVID-19 vaccine course during pregnancy: 28 who received Ad26.COV2.S (1 dose), 61 who received mRNA-1273 (2 doses), and 69 who received BNT162b2 (2 doses). Cohort demographics by vaccine platform are shown in Table 1. There was no significant difference in the number of days elapsed from the second dose of mRNA vaccines or single dose of Ad26.COV2.S vaccine to time of participant sample (median days [IQR]: 62 [27-91], 42 [22-74], and 58 [38-85] for mRNA-1273, BNT162b2 and Ad26.COV2.S respectively, p=0.10). There were no differences between study groups (vaccine platforms) in maternal age, gravidity, parity, pre-pregnancy BMI, race, insurance status, obesity, or presence of an autoimmune disorder. Individuals who received the Ad26.COV2.S vaccine were more likely to be of Hispanic ethnicity.

**Table 1.** Demographic and clinical characteristics of maternal participants by vaccine platform.

|  | Overall<br>(N=158) | Ad26.COV2.S<br>(N=28) | mRNA-1273<br>(N=61) | BNT162b2<br>(N=69) | *P* |
| --- | --- | --- | --- | --- | --- |
| Maternal age, years | 34 [32, 36] | 34 [32, 36] | 33 [32, 36] | 34 [32, 36] | 0.78 |
| Gravidity | 2 [1, 3] | 2 [2, 3] | 2 [1, 3] | 2 [1, 3] | 0.27 |
| Parity | 1 [0, 1] | 1 [0, 1] | 1 [0, 1] | 1 [0, 1] | 0.73 |
| Race (%) |  |  |  |  |  |
| Asian | 9 (6) | 1 (4) | 3 (5) | 5 (7) | 0.36 |
| Black or African American | 4 (3) | 2 (7) | 1 (2) | 1 (1) |  |
| Other | 6 (4) | 3 (11) | 2 (3) | 1 (1) |  |
| Unknown/Not Reported | 3 (2) | 1 (4) | 1 (2) | 1 (1) |  |
| White | 135 (85) | 21 (75) | 54 (89) | 60 (87) |  |
| American Indian or Alaskan Native | 1 (1) | 0 (0) | 0 (0) | 1 (1) |  |
| Ethnicity (%) |  |  |  |  |  |
| Hispanic | 9 (6) | 4 (14) | 3 (5) | 2 (3) | 0.02 |
| non-Hispanic | 141 (89) | 20 (71) | 57 (93) | 64 (93) |  |
| Unknown/Not reported | 8 (5) | 4 (14) | 1 (2) | 3 (4) |  |
| Insurance status (%) |  |  |  |  |  |
| Private | 152 (96) | 25 (89) | 61 (100) | 66 (96) | 0.05 |
| Public | 5 (3) | 3 (11) | 0 (0) | 2 (3) |  |
| Unknown | 1 (1) | 0 (0) | 0 (0) | 1 (1) |  |
| Pre-pregnancy BMI, kg/m<sup>2</sup> | 24 [22, 27] | 23 [22, 27] | 23 [22, 27] | 24 [22, 26] | 1.00 |
| Obesity (%) |  |  |  |  |  |
| No | 118 (75) | 22 (79) | 42 (69) | 54 (78) | 0.47 |
| Yes | 40 (25) | 6 (21) | 19 (31) | 15 (22) |  |
| Autoimmune condition (%) |  |  |  |  |  |
| No | 152 (96) | 28 (100) | 58 (95) | 66 (96) | 0.74 |
| Yes | 6 (4) | 0 (0) | 3 (5) | 3 (4) |  |
| Prior infection with SARS-CoV-2* (%) |  |  |  |  |  |
| No | 153 (97) | 26 (93) | 58 (95) | 69 (100) | 0.06 |
| Yes | 5 (3) | 2 (7) | 3 (5) | 0 (0) |  |

| Trimester of vaccination (%) |  |  |  |  |  |
| --- | --- | --- | --- | --- | --- |
| First | 18 (11) | 2 (7) | 7 (11) | 9 (13) | 0.05 |
| Second | 88 (56) | 13 (46) | 42 (69) | 33 (48) |  |
| Third | 52 (33) | 13 (46) | 12 (20) | 27 (39) |  |
| Time from vaccination to sample collection†, days | 50 [27, 86] | 58 [38, 85] | 62 [27, 91] | 42 [22, 74] | 0.10 |

Abbreviations: BMI, body mass index.
\*Four participants tested positive for SARS-CoV-2 prior to vaccination and one participant tested positive at delivery. †Defined time from single dose of Ad26.COV2.S vaccine or second dose of mRNA-1273 or BNT162b2 and sample collection. Continuous variables presented as median [IQR] and categorical variables as n (%). Differences between groups assessed by Kruskal-Wallis (continuous) or Fisher's exact test (categorical).

We initially evaluated transplacental antibody transfer via systems serology for those participants who had delivered at the time of maternal antibody profiling (n=123 maternal-neonatal dyads, Supplemental Table 1). There were no differences in gestational age at delivery, mode of delivery, neonatal sex, or neonatal birthweight by vaccine platform. To enhance understanding of transplacental antibody transfer by trimester of vaccination, IgG titers against Spike were quantified using ELISA in these 123 dyads and an additional 52 dyads who had delivered by study completion. In this set of 175 dyads, 27 participants (15%) were vaccinated with Ad26.COV2.S, 62 (35%) with mRNA-1273, and 86 (49%) with BNT162b2. Supplemental Table 2 depicts vaccine type and days elapsed from second dose (or single dose if receiving Ad26.COV2.S) to delivery by trimester of vaccination for this expanded dyad cohort.

### Maternal vaccine immune response by vaccine platform

To begin to understand differences in the vaccine-induced immune response across the three vaccine platforms, we plotted the Spike-specific antibody titer and Fc-receptor (FcR) binding in maternal serum (Figure 1A and Figure S1A). These plots reveal that whereas similar antibody profiles against Spike were observed for the two mRNA vaccines (mRNA-1273 and BNT162b2), vaccine-induced antibody titers and FcR-binding across all IgG subclasses and two antibody isotypes (IgG and IgA) were significantly lower in individuals who received the Ad26.COV2.S vaccine. Overall, the anti-Spike response was similar in individuals who received the mRNA vaccines mRNA-1273 and BNT162b2, with the exception of a significantly higher IgG2 anti-Spike response in women vaccinated with mRNA-1273 compared to those vaccinated with BNT162b2 (Figure S1A). Individuals who received Ad26.COV2.S also displayed significantly lower antibody functions (Figure 1B and S1C), as measured by antibody-dependent cellular phagocytosis (ADCP), antibody-dependent neutrophil phagocytosis (ADNP), antibody-dependent complement deposition (ADCD) and antibody-dependent NK-cell activation (ADNKA, measured as % CD107a+, % MIP-1β+ and % IFNγ+ cells). Antibody titer and FcR-binding against the Spikes from variants of concern (Alpha, Beta, Delta and Gamma) were highly correlated with response to the ancestral Spike, suggesting that individuals who mount a robust vaccine-induced antibody response will have antibodies against variants of concern (Figure S1B).

**Figure 1.**
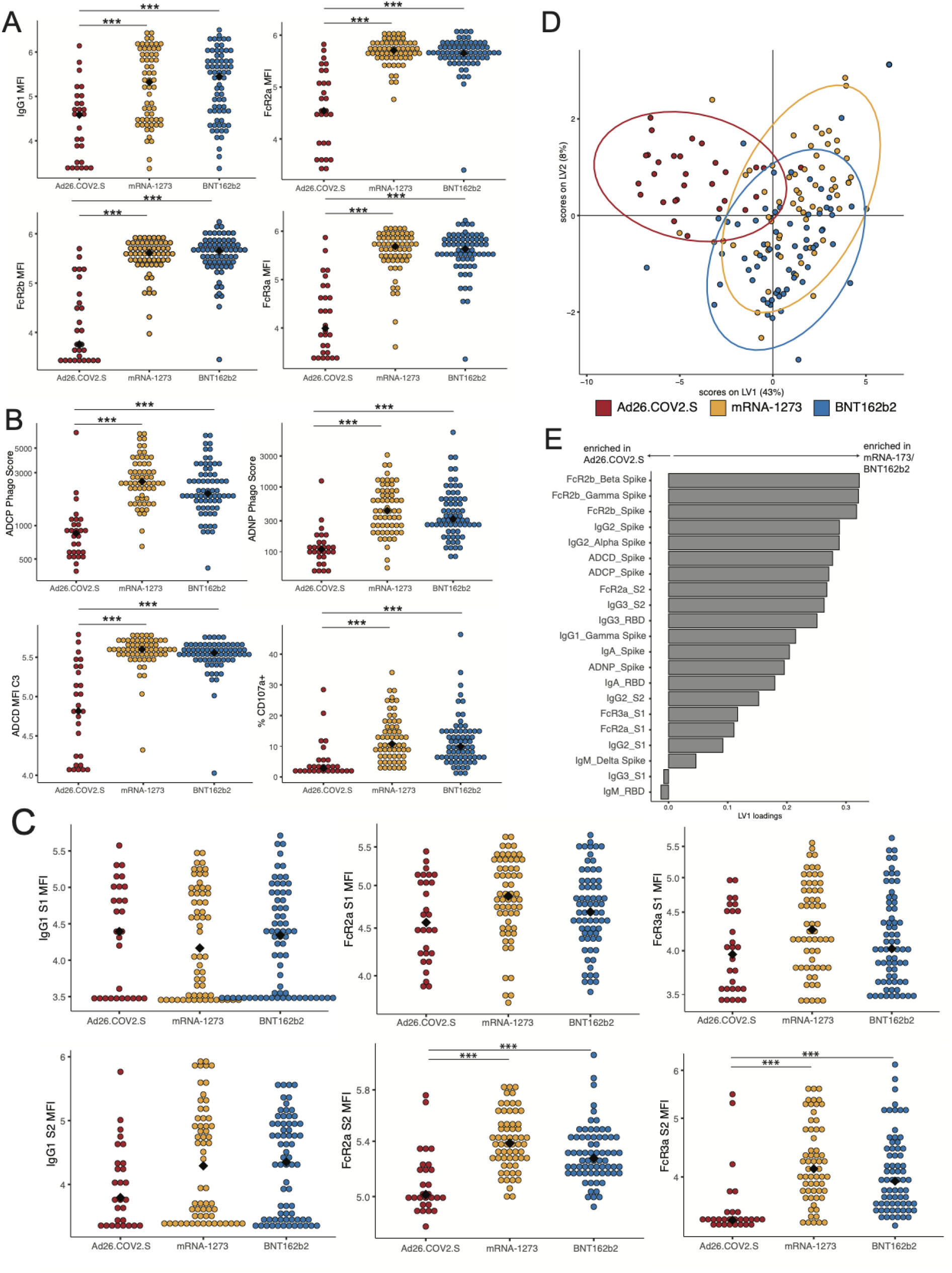
Maternal vaccine-induced titers are comparable between mRNA-1273 and BNT162b2 vaccination but lower after Ad26.COV2.S vaccination (A) Spike-specific IgG1 and Fc-receptor (FcR) binding were measured by Luminex. The dot plots show the titer for pregnant individuals who received Ad26.COV2.S (red), mRNA-1273 (yellow) or BNT162b2 (blue). (B) The dot plots show the Spike-specific antibody-dependent cellular phagocytosis (ADCP), antibody-dependent neutrophil phagocytosis (ADNP), antibody-dependent complement deposition (ADCD) and antibody-dependent NK cell degranulation, as measured by % CD107a + NK cells, in maternal samples. (C) The dot plots show the S1- (top) or S2- (bottom) specific IgG1 or FcR-binding in maternal samples. Significance was determined by Kruskal-Wallis test followed by posthoc Benjamini-Hochberg correction adjustment, * p < 0.05, ** p < 0.01, *** p < 0.001. (D) A partial-least squares discriminant model (PLSDA) was built using least absolute shrinkage and selection operator (LASSO)-selected SARS-CoV-2 specific antibody features in maternal samples, using vaccine type as the outcome variable. Each dot represents a sample, with the color representing the vaccine type. The ellipses represent the 95% confidence interval for the vaccine. (E) The barplot shows the latent variable (LV) 1 for the LASSO-selected features for the PLSDA in (D). Features with a positive loading along LV1 are enriched in mothers who received an mRNA vaccination, and features with a negative loading are enriched in mothers who received Ad26.COV2.S. Nearly all features are enriched in mRNA vaccine recipients.

To understand whether the different vaccine platforms elicited antibodies directed at different epitopes of Spike, we plotted the S1- and S2-specific IgG1 and FcR-binding in maternal plasma by vaccine. Interestingly, the IgG1 titer directed against S1 was comparable across vaccine platforms, whereas women who received Ad26.COV2.S had a nonsignificant decrease in IgG1 titer against S2 (Figure 1C). FcR-binding against the S1 domain was similar among vaccine platforms, whereas the FcR-binding against S2 was significantly lower for individuals who received Ad26.COV2.S (Figure 1C). These data suggest that differences in the FcR-binding of antibodies against S2 are primary drivers of reduced Ad26.COV2.S functions against Spike. Moreover, these data highlight that a single dose of Ad26.COV2.S can induce a similar S1-directed response as two doses of mRNA-1273 and BNT162b2.

To further examine differences in maternal vaccine response across vaccine platforms, a partial least squares discriminant analysis (PLSDA) was performed. Least absolute shrinkage and selection operator (LASSO) was used to select features most important to the model to prevent overfitting. This analysis revealed that although the vaccine responses in individuals who received mRNA-1273 or BNT162b2 were indistinguishable, the vaccine response in individuals who received Ad26.COV2.S was clearly separated from those who received either mRNA vaccine (Figure 1D). Nearly all LASSO-selected features were enriched in the women who received mRNA vaccination (Figure 1E).

### Maternal immune response by trimester of vaccination

We next sought to determine how the trimester of vaccination impacts the maternal vaccine-induced antibody response. Univariate analyses examining responses by trimester did not reveal trimester-specific differences in anti-Spike antibodies or FcR-binding (Figure 2A-B, Figure S2A-B). To further investigate the relative contribution of trimester of vaccination to anti-Spike antibody titer, FcR-binding, and function, the mean percentile rank of each feature was plotted by trimester of vaccination (Figure 2C). This analysis revealed that both first and third trimester vaccination drove a higher functional antibody response compared to second trimester vaccination, marked by both higher FcR-binding and more functional antibodies as indicated by enhanced ADCD, ADNP, ADCP, and ADNKA responses.

**Figure 2.**
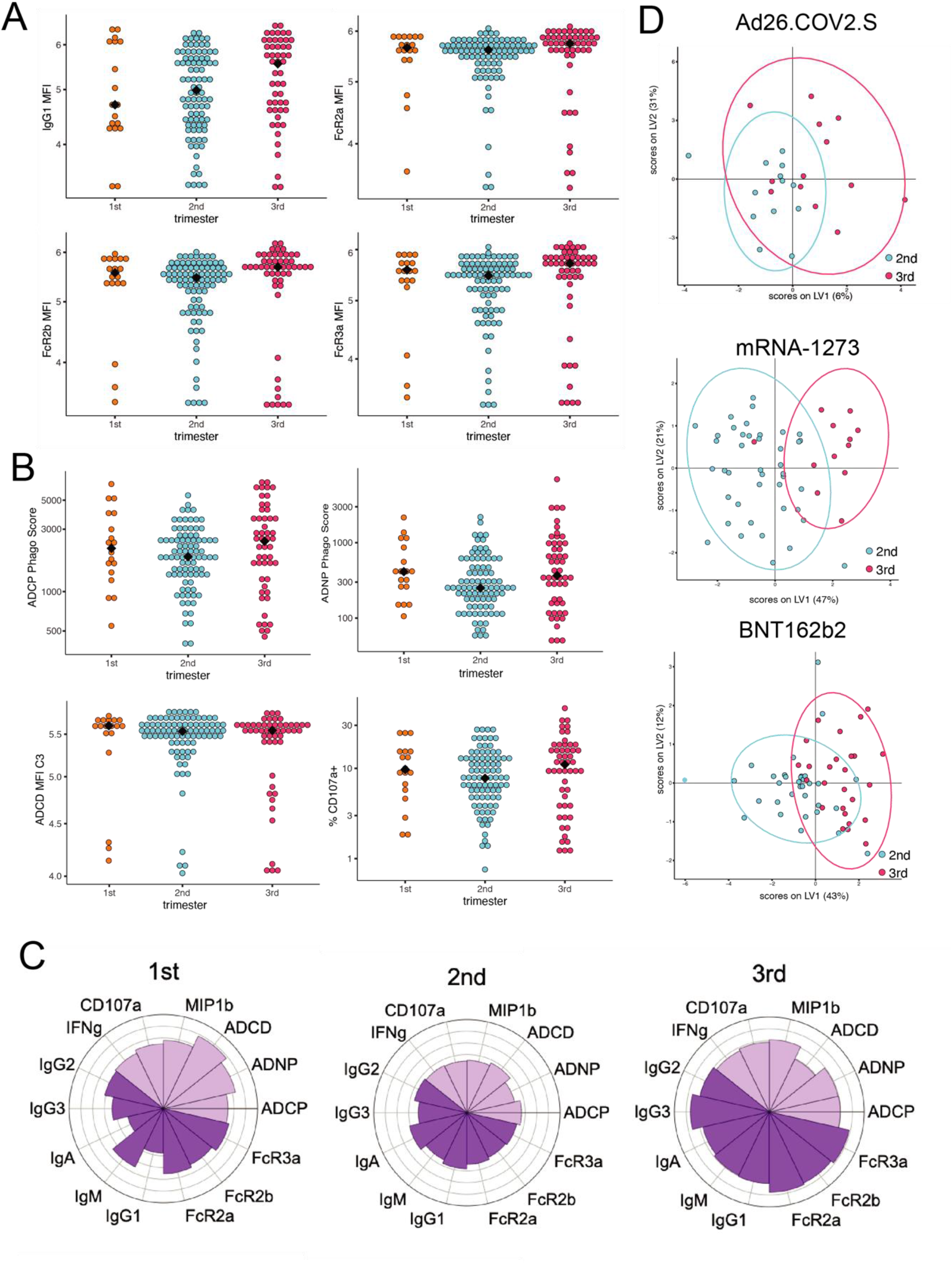
Trimester of vaccination affects vaccine-induced antibody titer in maternal samples. (A) The dot plots show the Spike-directed IgG1 and FcR-binding in maternal samples by trimester of vaccination. Significance was determined by Kruskal-Wallis test followed by posthoc Benjamini-Hochberg correction adjustment. No significant differences were found. (B) The dot plots show the Spike-directed antibody-dependent cellular phagocytosis (ADCP), antibody-dependent neutrophil phagocytosis (ADNP), antibody-dependent complement deposition (ADCD) and antibody-dependent NK cell degranulation, as measured by % CD107a + NK cells, in maternal samples by trimester of vaccination. Significance was determined by Kruskal-Wallis test followed by posthoc Benjamini-Hochberg correction adjustment. No significant differences were found. (C) The polar plots show the mean percentile rank for Spike-specific features in first, second, and third trimester of vaccination. (D-F) A PLSDA was built using LASSO-selected antibody features in maternal plasma for mothers who received Ad26.COV2.S (D), mRNA-1273 (E) or BNT162b2 (F) using trimester of vaccination as the outcome variable. Each dot represents a sample, with the color representing the trimester. The ellipses represent the 95% confidence interval for the trimester.

Given the observed differences in immune response driven by vaccine platform, we next sought to define the combination of features that best separate vaccine responses by trimester of vaccination within each vaccine platform group. To this end, LASSO was used to pick a minimal set of features that differentiated individuals vaccinated in the second and third trimesters, followed by PLSDA to visualize the separation between the second and third trimesters (Figure 2D); due to the smaller sample size, first trimester responses were not included in these analyses. Whereas there was little separation between second and third trimester vaccine responses in women that received the Ad26.COV2.S vaccine (5-fold CV: 0.3), there was a clear separation between the trimesters in women that received the mRNA-1273 vaccines (5-fold CV: 0.89, p < 0.05) and a modest separation between the trimesters in women that received the BNT162b2 vaccine (5-fold CV: 0.73, p < 0.05). The LASSO-selected features show an enrichment of antibody measurements in the third trimester relative to second within mRNA vaccine groups (Figure S2C-D). Specifically, individuals who received mRNA-1273 during the third trimester had an enrichment in IgA and IgG2 against variants of concern Alpha and Beta, and enrichment of ADCP compared to those who received mRNA-1273 in the second trimester (Figure S2C). This elevation in the IgA and IgG2 response in mRNA-1273 recipients was linked to a highly correlated response across SARS-CoV-2 variants (Figure S2C), and the increase in ADCP was strongly correlated with FcR-binding across variants of concern and ADNP activity in these women (S2C). Women who received BNT162b2 in the third trimester had enriched FcR2b-binding and IgM against the Alpha variant, and enriched IgG3 and ADNKA (measured by CD107a expression) responses compared to women who received BNT162b2 in the second trimester (Figure S2D). The increase in Alpha FcR2b-binding seen in third trimester BNT162b2 recipients was highly correlated with FcR-binding and IgG3 titer across SARS-CoV-2 variants, showing that these antibodies are highly inflammatory and likely highly functional.

### Transplacental antibody transfer by vaccine platform

To assess differences in the vaccine-induced immune response transferred from maternal to fetal circulation by vaccine type, we plotted the Spike-specific antibody titer and FcR-binding in umbilical cord serum in the 123 dyads who underwent systems serology profiling. In the cord blood, Spike-specific antibody titers and Fc-receptor binding were significantly higher in recipients of mRNA-1273 or BNT162b2 compared to recipients of Ad26.COV2.S (Figure 3A and Figure S3A). IgG2 against Spike was significantly higher in the cord blood of mRNA-1273 recipients compared to either Ad26.COV2.S or BNT162b2 recipients (Figure S3A). Moreover, Spike-specific antibody titers and functions in cord blood were highly correlated with the antibody response against variants of concern across all vaccine platforms, suggesting that Spike-specific antibodies in cord blood are likely to be active against variants of concern (Figure S3B). These observed differences in Spike-specific antibody response and response to variants of concern by vaccine platform in umbilical cord blood mirrored those observed in the maternal antibody response.

**Figure 3.**
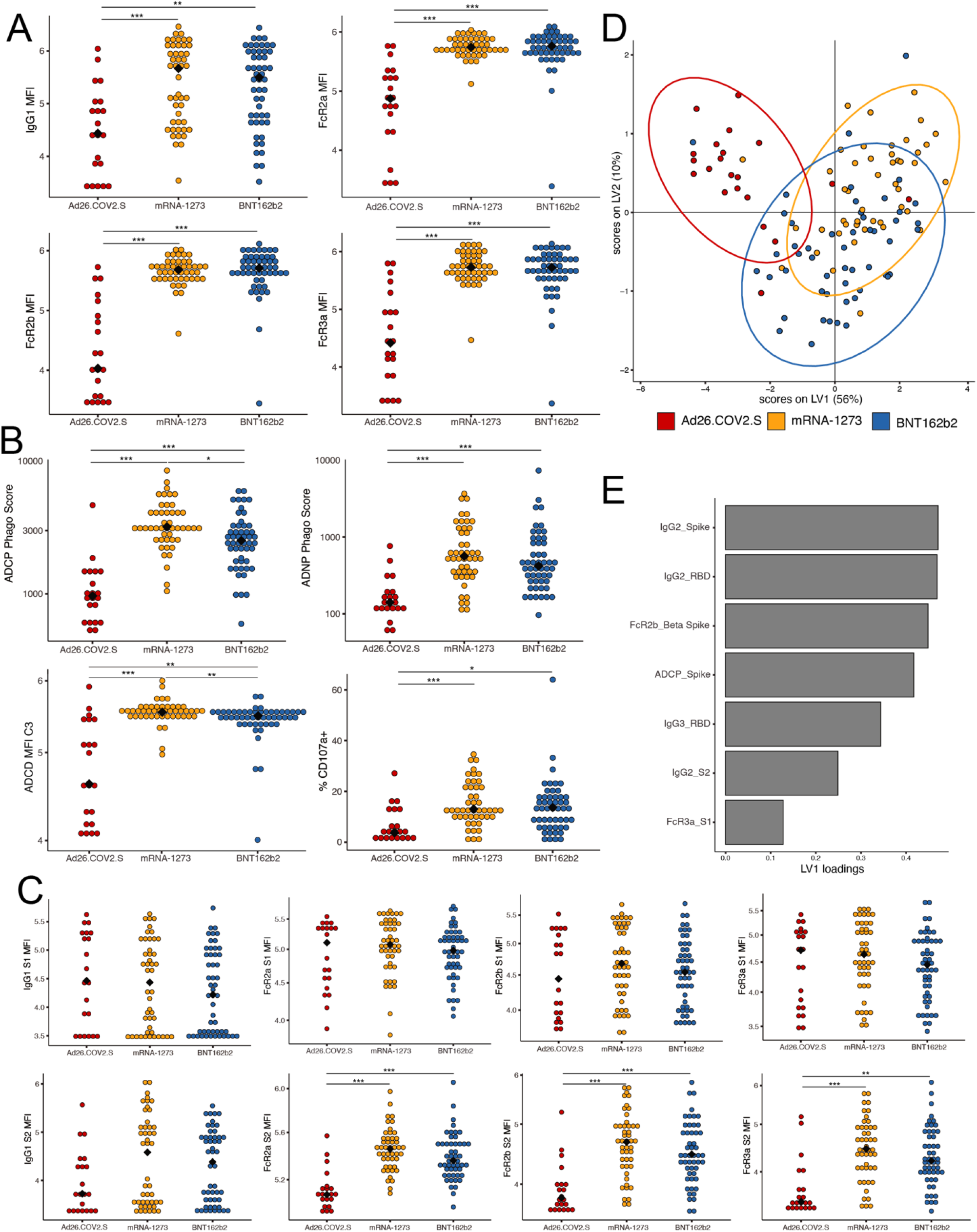
Vaccine-induced antibody titer in cord samples is comparable between mRNA-1273 and BNT162b2 vaccination but lower after Ad26.COV2.S vaccination (A) Spike-specific IgG1 and Fc-receptor (FcR) binding were measured by Luminex. The dot plots show the titer for cords whose mothers who received Ad26.COV2.S (red), mRNA-1273 (yellow) or BNT162b2 (blue). (B) The dot plots show the Spike-specific antibody-dependent cellular phagocytosis (ADCP), antibody-dependent neutrophil phagocytosis (ADNP), antibody-dependent complement deposition (ADCD) and antibody-dependent NK cell degranulation, as measured by % CD107a + NK cells, in cord samples. (C) The dot plots show the the S1 (top) or S2 (bottom) specific IgG1 or FcR-binding in cord samples. Significance was determined by Kruskal-Wallis test followed by posthoc Benjamini-Hochberg correction adjustment, * p < 0.05, ** p < 0.01, *** p < 0.001. (D) A partial-least squares discriminant model (PLSDA) was built using LASSO-selected SARS-CoV-2 specific antibody features in cord samples, using vaccine type as the outcome variable. Each dot represents a sample, with the color representing the vaccine type. The ellipses represent the 95% confidence interval for the vaccine. (E) The barplot shows the latent variable (LV) 1 for the least absolute shrinkage and selection operator (LASSO)-selected features for the PLSDA in (D). Features that with a positive LV1 loading were enriched in the cords whose mothers received an mRNA vaccine.

Functional antibody responses, including ADCP, ADNP, ADCD, and ADNKA (measured as % CD107a+, % MIP-1β+ and % IFNγ+ cells), were lower in the cord blood of Ad26.COV2.S recipients compared to mRNA-1273 or BNT162b2 (Figure 3B and S3C). Interestingly, the ADCP and ADCD responses in the cord blood of women who received mRNA-1273 were significantly higher than those of women who received either Ad26.COV2.S or BNT162b2 (Figure 3B), whereas the response in the maternal blood was similar between those two vaccines (Figure 1B and Figure S1B), suggesting preferential transfer of these highly-functional antibodies in mRNA-1273 recipients. Similar to what was observed in the maternal blood, vaccination with Ad26.COV2.S resulted in equivalent IgG1 titer and FcR-binding against S1 in cord blood compared to mRNA vaccination, but significantly lower FcR-binding antibodies (FcR2a, FcR2b, and FcR3a) against S2 in cord blood (Figure 3C).

A LASSO-PLSDA model was built using antibody features in cord blood to elucidate which antibody classes are enriched in the cord blood across vaccine platforms (Figure 3D). Whereas the mRNA-1273 and BNT162b2 cord blood responses had significant overlap, the Ad26.COV2.S cord blood response separated from the two other vaccine responses (Figure 3D). Moreover, all LASSO-selected features were enriched in the cord blood of women who had received mRNA-1273 or BNT162b2 compared to Ad26.COV2.S (Figure 3E). These data demonstrate strong similarities between maternal and cord blood antibody titers and functions, and reduced titer, FcR-binding and functionality of cord blood antibodies in recipients of Ad26.COV2.S relative to the mRNA vaccines.

To further probe the contribution of vaccine type on transplacental antibody transfer, we plotted the matched maternal-cord antibody titers and functions for each vaccine and compared differences (Figure 4A-B). While we expect titers to be higher in the cord relative to maternal serum for most vaccine-induced antibodies,^33–35^ it was notable that no antibody feature was significantly higher in the cord blood of women who had received the Ad26.COV2.S vaccine (Figure 4A-B). In contrast, nearly all Spike-specific antibody functions were higher in cord blood of women who received mRNA-1273 or BNT162b2 compared to maternal blood (ADCP, ADNP, and ADNKA by %CD107a+), with the exception of ADCD which was significantly *lower* in the cord relative to maternal blood of BNT162b2 recipients (Figure 4B).

**Figure 4.**
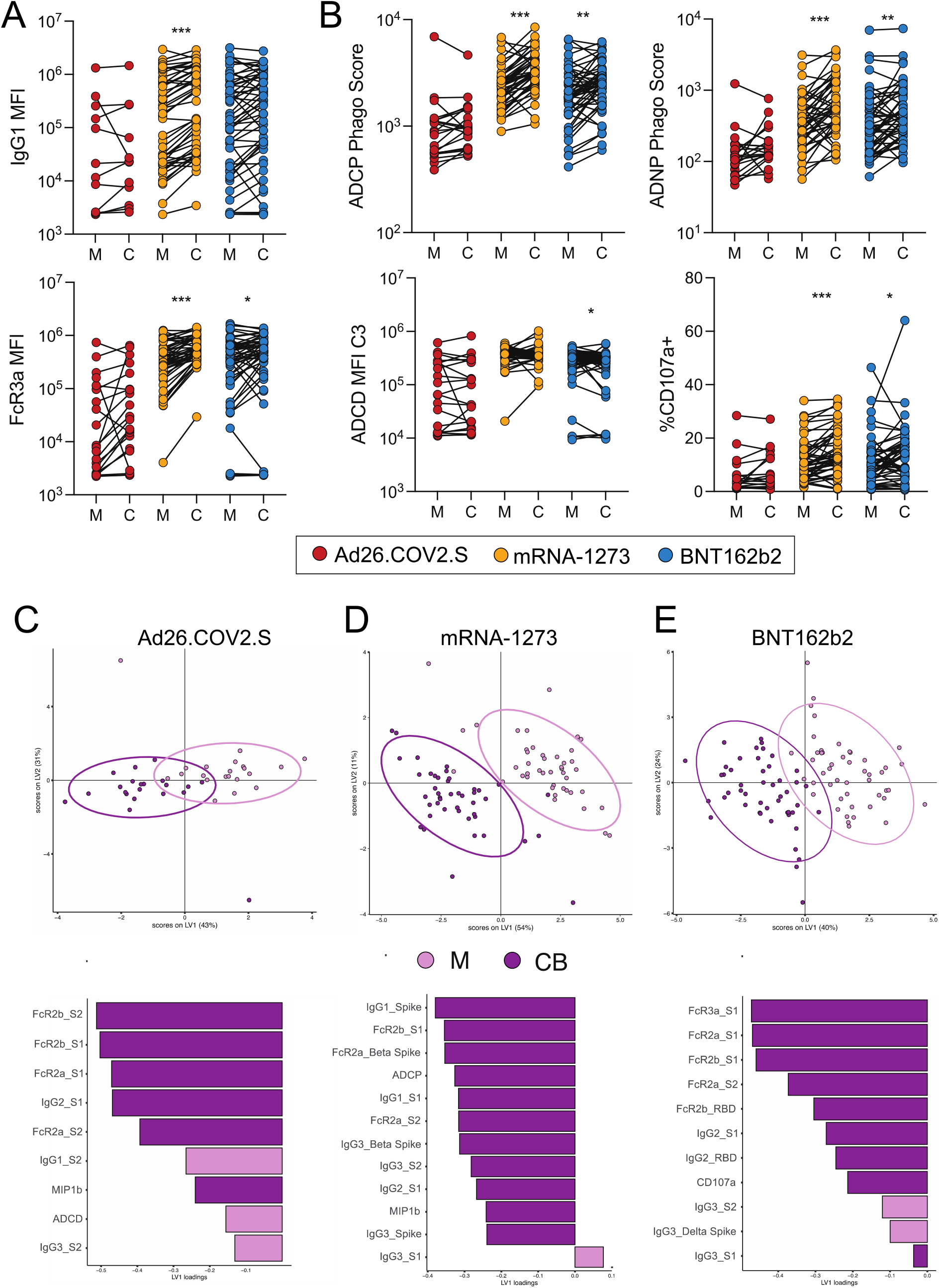
Efficient transfer of vaccine-induced antibodies to cord blood. (A) The dot plots show the Spike-specific IgG1 titer or FcR3a binding for maternal plasma (M) and cord blood (C). Lines connect maternal-cord dyads and the color represents vaccine type, Ad26.COV2.S (red), mRNA-1273 (yellow) or BNT162b2 (blue). Significance was determined by Wilcoxon signed rank test followed by posthoc Benjamini-Hochberg correction adjustment, * p < 0.05, ** p < 0.01, *** p < 0.001. (B) The dot plots show the Spike-specific antibody-dependent cellular phagocytosis (ADCP), antibody-dependent neutrophil phagocytosis (ADNP), antibody-dependent complement deposition (ADCD) and antibody-dependent NK cell degranulation, as measured by % CD107a + NK cells, for maternal plasma (M) and cord blood (C). Lines connect maternal-cord dyads and the color represents vaccine type, Ad26.COV2.S (red), mRNA-1273 (yellow) or BNT162b2 (blue). Significance was determined by Wilcoxon signed rank test followed by posthoc Benjamini-Hochberg correction adjustment, * p < 0.05, ** p < 0.01, *** p < 0.001. (C-E) A multilevel PLSDA (mPLSDA) was built for Ad26.COV2.S (C), mRNA-1273 (D) and BNT162b2 (E) using sample type, maternal blood (M, light purple) or cord blood (CB, dark purple) as the outcome variable. Features were selected using LASSO prior to building the models. The dot plots (top) show the scores plots for the mPLSDA. Each dot represents a sample, with the color representing the sample type. The ellipses represent the 95% confidence interval for the sample type. The bar plot show the LV1 for the mPLSDA built in each respective subfigure.

Given that transplacental transfer of antibody is driven substantially by maternal titers,^36,37^ it is possible that the lower transfer of antibodies in women that received Ad26.COV2.S could simply be due to lower maternal titers after Ad26.COV2.S compared to the mRNA vaccines. To reveal whether different vaccine platforms result in an enrichment of different antibody features in the cord blood, we performed a multilevel PLSDA (mPLSDA) using LASSO to select features that were most different between maternal and cord blood for each vaccine (Figure 4C-E, Figure S4). This approach accounts for the heterogeneous responses between vaccine recipients at the individual level. All three vaccines showed separation between maternal and cord blood and an enrichment of FcR-binding and Spike-specific IgG titer in the cord blood relative to maternal (Figure 4C-E, Figure S4). Interestingly, while we did not observe any significant differences between maternal and cord blood in Ad26.COV2.S vaccine recipients through univariate analysis, on a multivariate level, FcR2-binding antibodies, anti-S1 IgG2, and NK-cell-activating antibodies (MIP-1β) were enriched in cord blood of the Ad26.COV.S dyads, while maternal blood was enriched for anti-S2 IgG1 and IgG3, and ADCD (Figure 4C, Figure S4A). Thus, despite lower efficiency of antibody transfer in women that received Ad26.COV2.S, all three vaccines allowed for preferential transfer of specific antibodies to the cord blood. Similarly, the cord blood of mRNA-1273 or BNT162b2 recipients was enriched for IgG1 and IgG2, FcR-binding antibodies, functional antibodies, and antibodies directed against variants of concern (Alpha, Beta, and Delta), whereas maternal blood was enriched for IgG3 (Figure 4D-E, Figure S4B-C). These data highlight that although slight differences in transfer efficiency exist between vaccines, placental enrichment for highly functional antibody in the umbilical cord is a commonality that likely reflects a fundamental principle of transplacental transfer biology.

### Transplacental antibody transfer by trimester of vaccination

To investigate the impact of trimester of vaccination on the transplacental transfer of vaccine-induced immunity to the neonate at delivery, we measured total anti-Spike antibody IgG (as assessed by ELISA, see Methods) in the 123 dyads included in systems serology analyses and an additional 52 dyads who had delivered by study completion (N=175 total dyads, Supplemental Table 3). Interestingly, anti-Spike antibody titers in umbilical cord blood were higher than maternal titers at delivery when vaccination occurred in the first and second but not third trimesters (Figure 5A).

**Figure 5.**
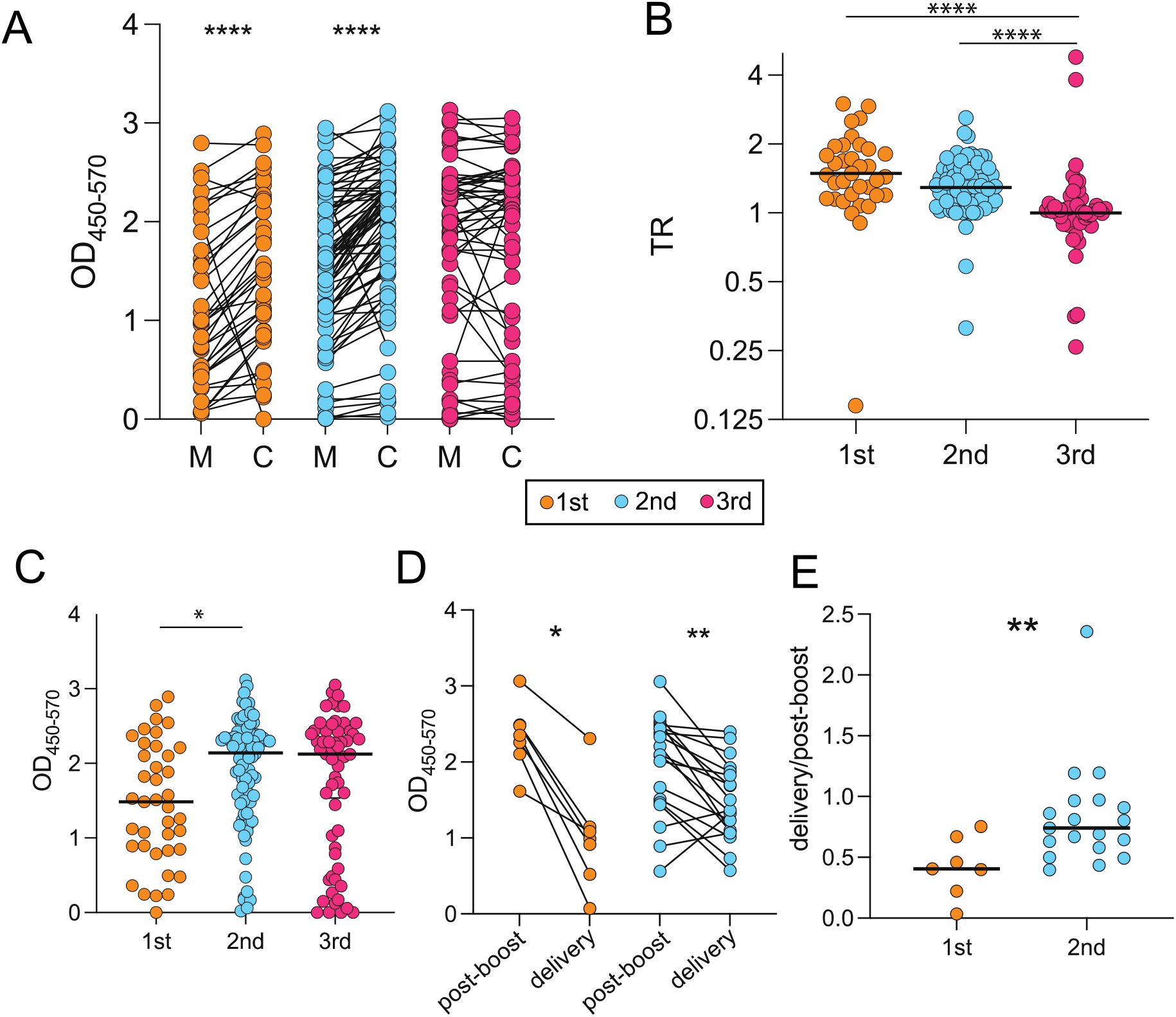
Transfer efficiency differs by trimester of vaccination A. The dot plots show the Spike-specific IgG titer in maternal (M) or cord (C) plasma. Lines connect paired dyads. The color indicates the trimester of vaccination, first (orange), second (blue) or third (pink). Significance was determined by a Wilcoxon signed rank test, * p < 0.05, ** p < 0.01, *** p < 0.001, **** p < 0.0001. B. The dot plot shows the transfer ratio (cord titer/maternal titer) of Spike-specific IgG to the cord. Color indicates trimester of vaccination. Significance was determined by Kruskal-Wallis test. **** p < 0.0001. C. The dot plot shows the Spike-specific IgG titer in cord blood by trimester that the mother received COVID-19 vaccination. Significance was determined by a one-way ANOVA followed by correction for multiple comparisons, * p < 0.05. D. The dot plots show the IgG Spike titer in maternal plasma post-boost (∼2-6 weeks after second dose of mRNA vaccine or after single dose of Ad26.COV2.S vaccine) and at delivery. Lines connect matched samples. Significance was determined by a Wilcoxon signed rank test, * p < 0.05, ** p < 0.01. E. The dot plot shows the ratio of the IgG Spike titer delivery/post-boost. Significance was determined by Mann Whitney test, ** p < 0.01.

The transfer ratio (TR) – defined as the ratio of cord blood anti-Spike IgG titer to maternal anti-Spike IgG titer at delivery – was calculated for each maternal-neonatal dyad and plotted by trimester of vaccination as a metric of transfer efficiency (Figure 5B). This analysis revealed higher TRs generated by first and second trimester vaccination (median TR = 1.5 and 1.3) compared to third trimester vaccination (median TR = 1.0). For reference, the expected efficiency of transplacental antibody transfer is > 1, indicating higher cord titers at delivery compared to maternal titers, with TRs of 1.2-3 at delivery noted for other vaccine-induced titers, such as measles, influenza, and pertussis.^33–35^

We next sought to determine absolute anti-Spike IgG titer in the cord blood at delivery by trimester of vaccination. Total anti-Spike IgG after first trimester vaccination was significantly lower than that in the cord blood of second trimester vaccine recipients (Figure 5C). In the context of the highest TRs observed in first trimester vaccine recipients, this finding likely reflects a waning of maternal titers at delivery compared to second trimester vaccine recipients. Given the finding of highest TRs for first trimester vaccination but lower absolute titers in cords of mothers vaccinated in the first trimester, suggestive of high placental efficiency in the setting of maternal first trimester vaccination but waning maternal titers by delivery, we next sought to quantify the waning of maternal antibody titers in first and second trimester vaccine recipients from completion of vaccination to delivery. In the subset of dyads in whom blood was drawn at 2-6 weeks following second vaccine dose in mRNA vaccine recipients or following the single Ad26.COV2.S dose, and again at delivery (n=7 first trimester, n = 19 second trimester), we compared total maternal anti-Spike IgG post-boost (second dose) and at delivery. This analysis demonstrated that antibody titers were significantly lower at delivery compared to shortly after the boost dose in both first and second trimester vaccine recipients (Figure 5D). As expected, the ratio of titers at delivery to post-boost was lower for first trimester vaccine recipients compared to second trimester vaccine recipients (Figure 5E), likely reflecting a more significant waning of antibody titer with time since vaccination.

## Discussion

Pregnancy is a unique immunological epoch, requiring complex and trimester-specific alterations in the maternal immune response to both protect the maternal-neonatal dyad and promote maternal tolerance of the semi-foreign fetal allograft.^22–24^ The COVID-19 pandemic revealed key deficits in our knowledge of both normal and challenged pregnancy immunity as well as the maternal response to vaccines. Although many vaccines can be safely administered in pregnancy, with seasonal influenza and tetanus-diphtheria-acellular pertussis (Tdap) routinely recommended for all pregnant individuals,^21,38^ observational studies of vaccination in pregnancy have generated only a limited understanding of how vaccine and pregnancy characteristics interact to impact vaccine-induced immune profiles.^20,39–42^ Here, we present evidence of both platform- and trimester-specific differences in the maternal immune response to the three COVID-19 vaccines available in the United States. We demonstrate that while functional spike-specific maternal antibodies are generated by all vaccine platforms, antibody titers and functionality profiles are enhanced in response to mRNA vaccines when compared with Ad26.COV2.S, and with mRNA-1273 demonstrating subtle functional advantages over BNT162b2. Vaccination in the first and third trimesters induced greater immunogenicity when compared with second trimester vaccination, with further analysis of mRNA vaccine recipients indicating comparatively less functionality against variants of concern in second trimester versus third trimester vaccination. Although total spike-specific antibody titers were lower in the umbilical cord at delivery following first trimester vaccination, we observed the most efficient transfer of highly functional antibodies from mother to umbilical cord following first trimester vaccination, likely reflective of the placenta working to preserve protection for the newborn in the setting of waning maternal titers over time. For unvaccinated individuals who become pregnant, receiving COVID-19 vaccination in the first trimester may therefore optimize benefit for mother and fetus by protecting against hospitalization, severe morbidity, and death due to COVID-19 over the greatest amount of time during pregnancy, without significantly compromising neonatal immune protection. Both maternal and neonatal immunity can be further enhanced by first trimester maternal vaccination, followed by boosting in the third trimester, with boosting 6 months post-mRNA vaccines and two months post-Ad26.COV2.S vaccine now recommended by the CDC, American College of Obstetricians and Gynecologists, and Society for Maternal-Fetal Medicine.^43–45^

Assessment of maternal and cord humoral immunity across vaccine platforms demonstrated significantly higher Spike-specific antibody titers, FcR-binding and functional antibodies induced by both mRNA COVID-19 vaccines compared to those induced by the Ad26.COV2.S vaccine. Although Ad26.COV2.S vaccination induces a lower antibody response than the mRNA vaccines in pregnant individuals as has been observed in non-pregnant individuals,^30,31,46,47^ the majority of participants who received Ad26.COV2.S in pregnancy still had detectable antibody titers and functions after a single vaccine dose. Recent studies have underscored the importance of central and effector memory T-cell responses in Ad26.COV2.S recipients^48,49^ and improved antibody coverage and neutralizing capacity against variants of concern over the eight months following single vaccination,^50^ suggesting that maturation of B cell responses occurs in Ad26.COV2.S recipients without boosting, and is an important driver of antibody protection over time. Thus, our assessment of antibody titers and functions in these Ad26.COV2.S recipients at a median of 44 days post vaccine may not capture the full depth and breadth of the immune response. The observed differences in maternal antibody quantity and quality between Ad26.COV2.S vaccine recipients and mRNA vaccine recipients might be a reflection of a one-versus two-dose regimen, rather than a reflection of an inferior response to the Ad-vectored vaccine platform itself. This concept is supported by recent data suggesting that the Ad26.COV2.S vaccine protection is enhanced in a two-dose regimen,^51,52^ with “booster” dose recommended any time two months or more after initial dose to enhance protection in specific vulnerable populations.^43^ Further evaluation of whether the differences seen between mRNA vaccines and the Ad26.COV2.S vaccine persist after two doses of the Ad-vectored vaccine is a critical area for future research, and will elucidate whether the differences noted in pregnancy relate simply to dosing and interval, versus the Ad-vectored platform itself. As previous work has shown the importance of adherence to the prime/boost timeline for mRNA vaccine recipients given delayed kinetics of antibody responses during pregnancy,^13^ investigating the impact of “booster” doses given during pregnancy, particularly in recipients who originally received the Ad26.COV2.S vaccine, will be important to obtaining a full understanding of how vaccine strategies can be tailored to optimize maternal protection.

Robust changes in the inflammatory profile occur during pregnancy to facilitate implantation and early placentation, followed by a period of rapid fetal growth, and finally, the onset of parturition.^22–25^ How these immune fluctuations influence maternal responses to vaccines administered across gestation is not known. Although univariate analyses examining responses by trimester did not reveal trimester-specific differences, these analyses likely fail to account for interactions between multiple elements of the antibody response. Harnessing the strength of the systems serology approach, we identified superior immunogenicity – as characterized by anti-Spike antibody FcR-binding capacity and functionality – in pregnant individuals vaccinated in the first and third trimesters compared to those vaccinated in the second trimester through multivariate modeling. These data demonstrate the importance of considering more than just Ig titer alone when evaluating vaccine immunogenicity. Further investigation into the anti-Spike antibody responses (including against variants of concern) in second and third trimester vaccine recipients by vaccine type revealed that second trimester responses are impaired compared with third trimester for both mRNA vaccines. Taken together, these data suggest that second trimester vaccination generates an antibody response characterized by overall reduced FcR-binding capacity and functionality relative to vaccination in the first and third trimesters. These findings can be understood in the context of immunomodulatory changes that occur in the second trimester of pregnancy that favor maternal tolerance of the developing fetal semi-allograft and promote a state of immunological quiescence,^25^ in which response to non-self antigens may be dampened.

While effective maternal protection is paramount during a pandemic, neonatal protection against potentially harmful pathogens through maternal immunization is an important secondary consideration when developing vaccine recommendations in pregnancy.^20,21^ Recipients of all 3 COVID-19 vaccines demonstrated enrichment of FcR-binding and anti-Spike IgG titer in the cord compared with maternal blood, with the most favorable cord:maternal transfer ratios (greater than 1) following first and second trimester vaccination. Whether higher antibody transfer ratios observed for first trimester COVID-19 vaccination are due to differences in antibody Fc-quality and thus affinity for Fc-receptors that traffic antibody to the fetal circulation,^37,53,54^ or are the result of increased time for antibody transit to occur, or a combination of both is yet to be determined. As expected, the waning of maternal titers was more significant after first compared to second trimester maternal vaccination, and both maternal and neonatal immunity would therefore be boosted by maternal COVID-19 boosting in the third trimester, when initial vaccination (or vaccination series) occurs in the first trimester.

These results add significantly to our current understanding of how timing of maternal vaccination impacts both maternal immune response and transplacental transfer efficiency. Current clinical recommendations governing the timing of routinely administered vaccines in pregnancy (e.g. influenza vaccine, which is administered during influenza season regardless of trimester, and Tdap, which is administered in late second to early third trimester with the primary goal of enhancing transplacental transfer) have limited the ability to systematically investigate the impact of vaccine administration across gestation. The Advisory Committee on Immunization Practices (ACIP) has advised routine Tdap administration during pregnancy after 20 weeks of gestation for over a decade, based on limited availability of safety data in the first trimester.^55^ Studies that have investigated the timing of Tdap vaccination after the first trimester have found superior transfer of anti-pertussis antibody when vaccination occurs earlier in the recommended interval of 27-36 weeks,^41,56,57^ with some evidence from individuals vaccinated outside that window favoring improved transfer following early second trimester vaccination.^58^ Data on placental transfer of anti-pertussis antibody following Tdap administration in the first trimester are not available, as studies including first trimester vaccinees are primarily limited to safety reports.^59,60^ Data from seasonal influenza vaccine administration in pregnancy, which is administered at any gestational age during influenza season, are conflicting with respect to maternal immune response. Some studies suggest lower maternal anti-influenza titers in first compared to second trimester vaccination, with highest anti-influenza titers in third trimester vaccination,^42,61,62^ while others suggest a more robust maternal titer generated by first and third trimester vaccination relative to second.^63^ In addition, a majority of studies noted enhanced cord blood antibody titers against influenza following third trimester vaccination when compared with second or first trimester vaccination, likely due in part to waning maternal antibody titers with increased time from vaccination.^42^ These studies were limited by their narrow focus on IgG titer as the primary measure of maternal immune response, while our systems serology approach permits the dissection of diverse components of maternal humoral immunity. Whether differences in cord blood titers observed between first, second, and third trimester vaccination correlate with differential neonatal protection from COVID-19 will be important to assess in neonatal cohorts.

The rapid development and distribution of three novel COVID-19 vaccines in the US has offered an unprecedented opportunity to further our understanding of the rules of vaccine-induced immunity in pregnancy. Our study contributes substantially to knowledge of how the maternal-neonatal dyad responds to vaccination against a *de novo* antigen with novel mRNA and Ad-vectored vaccines, which were not specifically designed to optimize maternal or neonatal protection, as pregnant individuals were excluded from initial vaccine clinical trials.^9–11^ Looking beyond responses to the COVID-19 vaccines, our findings may have broader implications.

These insights into both platform- and trimester-specific differences to COVID-19 vaccines can be used to guide rational vaccine development and enhance understanding of the maternal immune response to varied perturbations across trimesters. Optimizing both maternal and neonatal immunity is a key consideration informing vaccination strategies in this unique population, and our data suggest that for the COVID-19 vaccines, maternal immunization in the first trimester may optimize benefit to both mother and neonate, particularly if boosting is employed in the third trimester. Efforts to recruit and include pregnant individuals in vaccine studies will remain critical to constructing evidence-based vaccine strategies that maximize benefit to both mother and newborn.

## Methods

### Participant recruitment and study design

Pregnant individuals at two tertiary care centers were approached for enrollment in the COVID-19 pregnancy biorepository study between January 2021 and September 2021, Protocol #2020P003538, approved by Mass General Brigham Institutional Review Board (IRB). Eligible participants were pregnant, greater than or equal to 18 years old, able to provide informed consent, and receiving the Ad26.COV2.S, mRNA-1273, or BNT162b2 COVID-19 vaccine during pregnancy. Eligible participants were identified by practitioners at the participating hospitals or were self-referred. A study questionnaire was administered to assess pregnancy status, history of prior SARS-CoV-2 infection, timing of COVID-19 vaccine doses, and type of COVID-19 vaccine received. Individuals were grouped by the type of vaccine received, and by the trimester at which the first vaccine dose was given in recipients of mRNA-1273 and BNT162b2 vaccines (or at the time of the single dose in recipients of the Ad26.COV2.S vaccine). To maximize the generalizability of our findings to the general pregnant population, participants who tested positive for SARS-CoV-2 prior to or after receiving a COVID-19 vaccine were not excluded from this study.

### Sample collection

For Ad26.COV2.S vaccine recipients, blood was collected at least 2 weeks after receiving the single vaccine dose. For mRNA-1273 and BNT162b2 vaccine recipients, blood was collected at least 2 weeks following the second vaccine dose. For participants who delivered during the study time frame (N=123), maternal blood was drawn at the time of delivery, and umbilical cord blood was collected after delivery. Blood was collected by venipuncture (or from the umbilical vein following delivery) into serum separator and EDTA tubes and centrifuged at 1000 g for 10 min at room temperature. Serum and plasma were aliquoted into cryogenic vials and stored at - 80°C.

### Antigens

Antigens used for assays included SARS-CoV-2 D614G Spike, Alpha Spike, Beta Spike, Gamma Spike and Delta Spike (all Spikes kindly provided by Erica Ollman Saphire) and SARS-CoV-2 S1 and S2 (Sino Biological).

### Primary Cells

Neutrophils were isolated from fresh peripheral whole blood collected at the Ragon Institute. NK cells were isolated from fresh peripheral blood from buffy coats collected at Massachusetts General Hospital (MGH). All volunteers gave signed, informed consent and were over the age of 18, and samples were deidentified before use. The study was approved by the MGH Institutional Review Board. Neutrophils were maintained in R10 media (RPMI supplemented 10% fetal bovine serum (FBS) (Sigma Aldrich), 5% penicillin/streptomycin (Corning, 50 μg/mL), 5% L-glutamine (Corning, 4 mM), 5% HEPES buffer (pH 7.2) (Corning, 50 mM)) and at 37°C, 5% CO2 for the duration of the assay. After isolation, NK cells were rested overnight at R10 media supplemented with 2ng/mL interleukin (IL)-15 at 37°C, 5% CO2.

### Bead-based functional assays

For antibody-dependent cellular phagocytosis (ADCP), antibody-dependent neutrophil phagocytosis (ADNP) and antibody-dependent complement deposition (ADCD), D614G Spike was biotinylated using Sulfo-NHS-LC-LC biotin (Thermo Fisher Scientific) and desalted using Zeba Columns (Thermo Fisher Scientific). Biotinylated antigen was coupled to yellow-green FluoSpheres NeutrAvidin beads (for ADCP and ADNP) or red neutravidin beads (for ADCD) (Invitrogen). To form immune complexes, antigen-coupled beads were incubated with appropriately diluted serum (1:100 for ADCP, 1:50 for ADNP and 1:10 for ADCD) for 2 hours at 37°C. Immune complexes were then washed. For ADCP, THP-1 cells (ATCC) were added to plates at a concentration of 2.5×10^4^ cells/mL. Cells were incubated for 16-18 hours at 37°C with the immune complexes and fixed following the incubation. Fluorescence was acquired using an iQue (Intellicyt). For ADNP, leukocytes were isolated from fresh peripheral blood using ACK Lysing Buffer (Thermo Fisher Scientific). Leukocytes were added at a concentration of 5×10^4^ cells/mL. Cells were incubated for 1 hour at 37°C with the immune complexes. Following the incubation, neutrophils were stained using anti-CD66b Pacblue (biolegend). Cells were then fixed. Fluorescence of CD66b+ cells was acquired using an iQue (Intellicyt). For ADCP and ADNP, a phago score was calculated using the following formula: (% fluorescent cells*MFI of fluorescent cells)/10000. For ADCD, lyophilized guinea pig complement (Cedarlane) was diluted in gelatin veronal buffer supplemented with calcium and magnesium. The diluted guinea pig complement was added to immune complexes and plates were incubated at 37°C for 20 minutes. Plates were washed with 15 μM EDTA diluted in PBS. Complement was stained using anti-C3 FITC (MP BioProducts). Fluorescence was determined using an iQue (Intellicyt). For all functional assays, samples were run in duplicate and data is reported as the average of the replicates.

### Antibody-dependent NK cell degranulation

ELISA plates were coated with 2 μg/mL of Spike protein. Plates were washed and blocked with 5% BSA in PBS. Immune complexes were formed by adding serum diluted 1:25 to plates and incubating plates for 2 hours at 37°C. RosetteSep (STEMCELL Technologies) and a ficoll gradient was used to isolate NK cells from fresh peripheral blood from healthy donors. Isolated NK cells were rested overnight in R10 (see *Primary Cells* section above) with 2 ng/mL of IL-15. NK cells were added to immune complexes at a concentration of 5 × 10^4^ cells/mL in media supplemented with Brefeldin A (Sigma), anti-CD107a BV605 (BD Biosciences) and GolgiStop (BD Biosciences). NK cells were incubated with immune complexes for 5 hours at 37°C. After incubation, cells were stained for surface markers using anti-CD56 PE-Cy7 (BD Biosciences) and anti-CD3 APC-Cy7 (BD Biosciences). Cells were fixed with PermA (Life Technologies), Permeabilized with Perm B (Life Technologies), and stained with anti-MIP1b-BV421 (BD Biosciences) and anti-IFNg-PE (BD Biosciences). The cells were analyzed for fluorescence using an iQue (Intellicyt). NK cells were gates as CD3-/CD56+ and NK activity was determined as percent of cells positive for CD107a, IFN-g or MIP-1b.

### Multiplexed Luminex Assay

A multiplexed Luminex assay was used to determine the relative concentration of antigen-specific antibody isotype and subclass titer and Fc receptor binding. Carboxylated microsphere were coupled to antigen using EDC and Sulfo-NHS (Thermo Fisher Scientific) to form covalent NHS-ester linkages. To form immune complexes, diluted serum (1:100 for IgG2/3, 1:500 for IgG1, and 1:1000 for FcRs) was mixed with antigen-couple microspheres and incubated overnight at 4°C shaking at 700 rpm. The following day, plates were washed three times with 0.1% BSA 0.02% Tween-20 in PBS. Antigen-specific antibody isotypes were measured using PE-coupled mouse anti-human antibodies (Southern Biotech) specific for each specific isotype. Avi-tagged FcRs (Duke Human Vaccine Institute) were biotinylated using a BirA500 kit (Avidity) and tagged with streptavidin-PE. PE-tagged FcRs were added to immune complexes to determine antigen-specific FcR binding. Fluorescence was acquired using an iQue (Intellicyt) and antigen-specific antibody titer and FcR-binding is reported as Median Fluorescence Intensity (MFI).

### Enzyme-linked immunosorbent assay (ELISA)

To further assess cord:maternal transfer ratios by trimester of vaccination, maternal and umbilical cord blood samples were collected from an additional 52 participants who delivered during the study period (N=175 total maternal-neonatal dyads included for this analysis). Antibodies against the SARS-CoV-2 Spike were quantified using an ELISA. ELISA plates were coated with 500 ng/mL of D614G Spike (kindly provided by Erica Saphire) and incubated for 30 minutes at room temperature. Plates were washed with washing buffer (0.05% Tween-20. 400mM NaCl, 50mM Tris, pH 8.0) and blocked with a 0.1% BSA solution for 30 minutes at room temperature. Plates were washed, and sample was added at a dilution of 1:100. Plates were incubated with sample at 37°C for 30 minutes. Plates were washed, and a horseradish peroxidase (HRP)-conjugated goat anti-human IgG antibody (Bethyl Laboratories) was added for detection of Spike-specific IgG. Plates were incubated with secondary antibody for 30 minutes at room temperature and then washed. TMB was used to develop the ELISA and sulfuric acid was used to stop the ELISA. Signal was read at 450 nm and background corrected from a reference wavelength of 570 nm.

### Statistical analysis

For univariate analysis, statistics were calculated using GraphPad Prism version 8.0. Luminex data and ADCD were log_10_-transformed before analysis. For analysis of differences between vaccines or trimesters, significance was determined by a one-way ANOVA followed by posthoc Benjamini-Hochberg adjustment. For analysis of maternal-cord differences, significance was determined by a Wilcoxon matched pairs signed rank test followed by posthoc Benjamini-Hochberg adjustment. Multivariate analysis was performed in R (version 4.0.0). Prior to building the models, data was centered and scaled. LASSO feature selection was performed using the “select_lasso” function in systemseRology R package (v1.0) (https://github.com/LoosC/systemsseRology) to determine significant features. The LASSO tuning parameter was determined by 5-fold cross validation. LASSO feature selection was performed 100 times, and features that were chosen 50% of the repetitions were selected to build the model. LASSO-selected features were used to build partial least squares discriminant analysis (PLSDA) or multilevel PLSDA models. Model performance was determined by 5-fold cross validation.

## Data Availability

All data produced in the present study are available upon reasonable request to the authors.

## Notes

### Competing Interest Statement

K.J.G. has consulted for Illumina, BillionToOne, and Aetion outside the scope of the submitted work. G.A. is the founder of Seromyx Inc. M.A.E. reported serving as medical advisor for Mirvie. A.F. reported serving as a cofounder of and owning stock in Alba Therapeutics and serving on scientific advisory boards for NextCure and Viome outside the submitted work. All other authors report no competing interests.

### Funding Statement

This study was funded by grants from NIH, March of Dimes, Gates Foundation and MassCPR to the authors, as described in the manuscript.

### Author Declarations

IRB of MassGeneral Brigham gave ethical approval for this work.

