## Supplemental File 1 for "Maternal immune response and placental antibody transfer after COVID-19 vaccination across trimester and platforms"

Supplemental Information

Supplemental Table 1. Pregnancy characteristics of systems serology maternal:neonatal dyads, by vaccine platform.

| | Overall (N=123) | Ad26.COV2.S (N=21) | mRNA-1273 (N=47) | BNT162b2 (N=55) | *P* |
|---|---|---|---|---|---|
| Gestational age at delivery, completed weeks | 39 [38, 40] | 39 [39, 40] | 39 [38, 40] | 39 [39, 40] | 0.57 |
| Mode of delivery (%) | | | | | |
| Vaginal delivery | 88 (72) | 13 (62) | 36 (77) | 39 (71) | 0.45 |
| Cesarean section | 35 (28) | 8 (38) | 11 (23) | 16 (29) | |
| Neonatal sex (%) | | | | | |
| Male | 73 (59) | 11 (52) | 28 (60) | 34 (62) | 0.76 |
| Female | 50 (41) | 10 (48) | 19 (40) | 21 (38) | |
| Neonatal birthweight, g | 3362 [3094, 3741] | 3390 [3155, 3820] | 3345 [3024, 3730] | 3395 [3141, 3716] | 0.90 |
| Prior infection with SARS-CoV-2* (%) | | | | | |
| No | 119 (97) | 19 (90) | 45 (96) | 55 (100) | 0.06 |
| Yes | 4 (3) | 2 (10) | 2 (4) | 0 (0) | |
| Trimester of vaccination (%) | | | | | |
| First | 3 (2) | 0 (0) | 3 (6) | 0 (0) | <0.001 |
| Second | 70 (57) | 9 (43) | 36 (77) | 25 (45) | |
| Third | 50 (41) | 12 (57) | 8 (17) | 30 (55) | |
| Time from vaccination to delivery†, days | 74 [38, 100] | 75 [52, 95] | 82 [61, 108] | 50 [20, 78] | 0.002 |

*Three participants tested positive for SARS-CoV-2 prior to vaccination and one participant tested positive at delivery †Defined time from single dose of Ad26.COV2.S vaccine or second dose of mRNA-1273 or BNT162b2 and sample collection at delivery. Continuous variables presented as median [IQR] and categorical variables as n (%). Differences between groups assessed by Kruskal-Wallis (continuous) or Fisher's exact test (categorical).

Supplemental Table 2. Characteristics of maternal:neonatal dyad cohort for transplacental antibody transfer ELISA analysis by trimester of vaccination.

| | All (N=175) | First (N=39) | Second (N=79) | Third (N=57) | *P* |
|---|---|---|---|---|---|
| Vaccine type (%) | | | | | |
| Ad26.COV2.S | 27 (15) | 3 (8) | 12 (15) | 12 (21) | <0.001 |
| mRNA-1273 | 62 (35) | 14 (36) | 40 (51) | 8 (14) | |
| BNT162b2 | 86 (49) | 22 (56) | 27 (34) | 37 (65) | |
| Prior infection with SARS-CoV-2* (%) | | | | | |
| No | 164 (94) | 34 (87) | 75 (95) | 55 (96) | 0.22 |
| Yes | 11 (6) | 5 (13) | 4 (5) | 2 (4) | |
| Time from vaccination to delivery†, days | 82 [45, 127] | 167 [156, 177] | 92 [74, 109] | 28 [19, 43] | <0.001 |

*Three participants tested positive for SARS-CoV-2 prior to vaccination and one participant tested positive at delivery †Defined time from single dose of Ad26.COV2.S vaccine or second dose of mRNA-1273 or BNT162b2 and sample collection at delivery. Continuous variables presented as median [IQR] and categorical variables as n (%). Differences between groups assessed by Kruskal-Wallis (continuous) or Fisher's exact test (categorical).

### Supplemental Figures

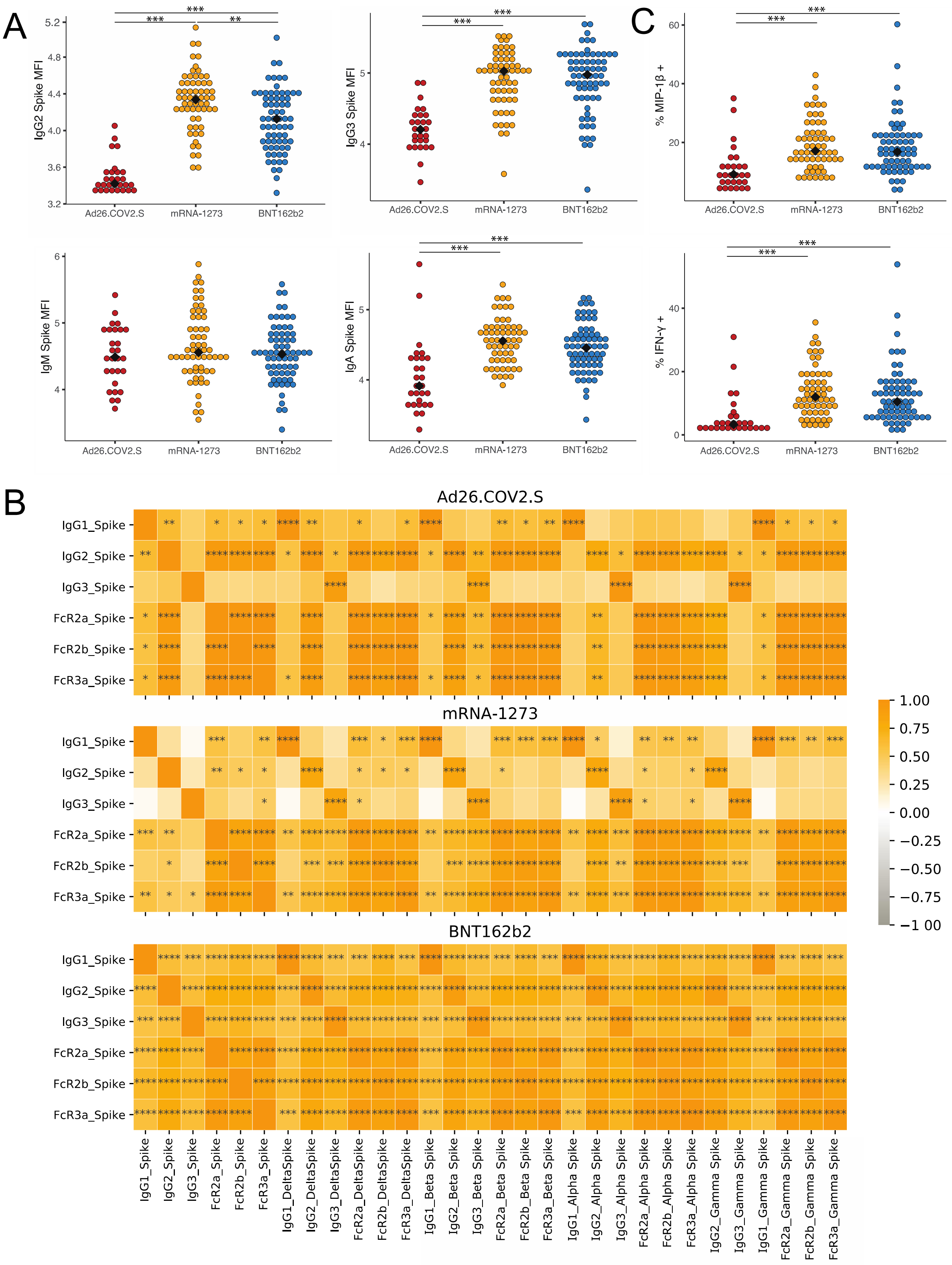

Figure S1. Vaccine type results in different vaccine-induced antibody induction in maternal samples

(A) Spike-specific IgG2, IgG3, IgM and IgA were measured by Luminex. The dot plots show the titer for mothers who received Ad26.COV2.S (red), mRNA-1273 (yellow) or BNT162b2 (blue).

(B) The heatmaps show the spearman correlation of antibody features against the Spike of the ancestral virus versus antibody features against the Spikes from variants of concern in cord blood, analyzed separately for each vaccine platform. Orange indicates a positive spearman correlation, * $p < 0.05$, ** $p < 0.01$, *** $p < 0.001$, **** $p < 0.0001$

(C) The dot plots show the Spike-specific antibody-dependent NK cell activation, as measured by % MIP-1b + and % IFN-g+ NK cells, in maternal blood. Significance was determined by Kruskal-Wallis test followed by posthoc Benjamini-Hochberg correction adjustment, * $p < 0.05$, ** $p < 0.01$, *** $p < 0.001$

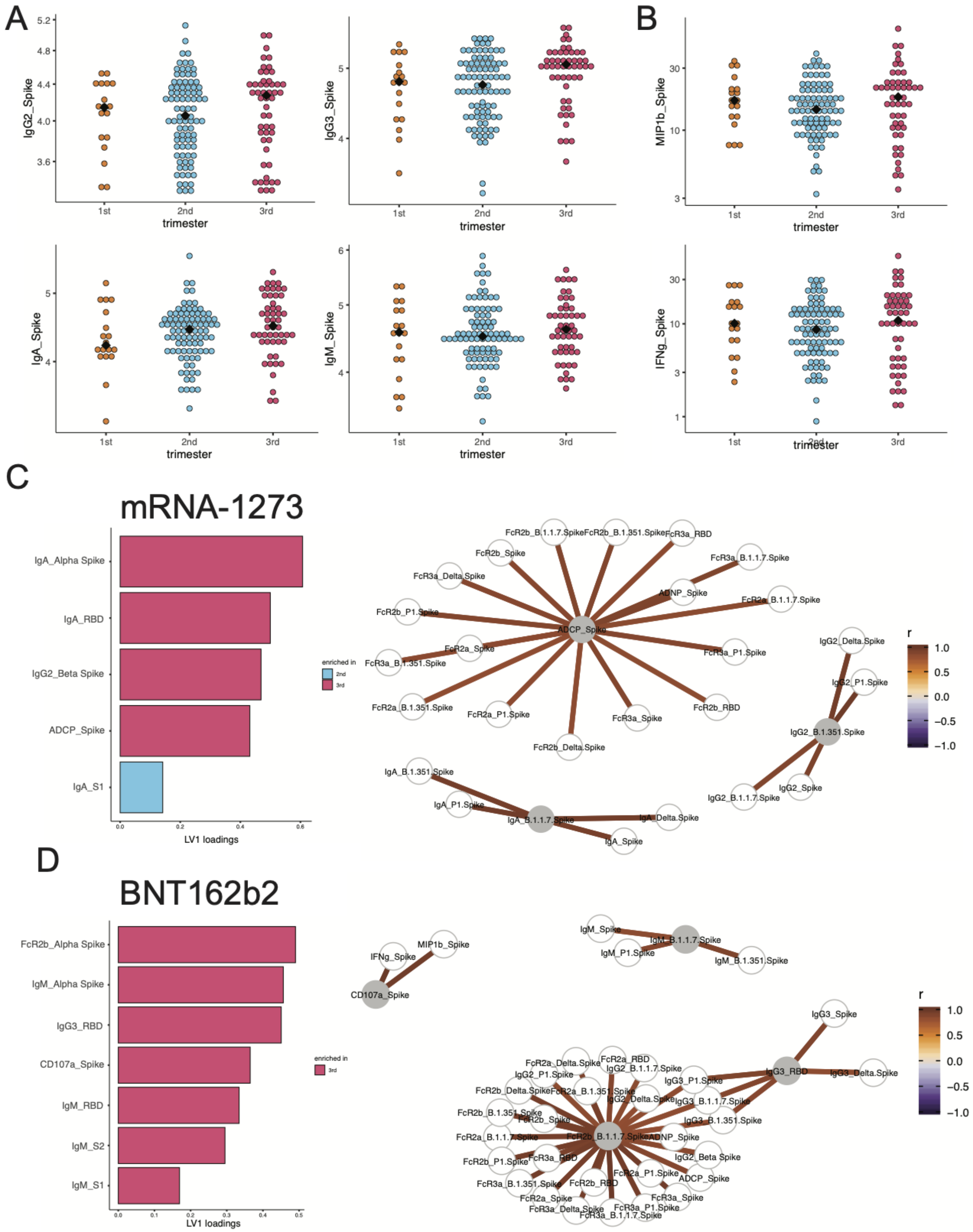
A
B
C
D
IgG2_Spike
IgG3_Spike
MIP1b_Spike
IgA_Spike
IgM_Spike
IFNg_Spike
trimester
1st
2nd
3rd
mRNA-1273
IgA_Alpha Spike
IgA_RBD
IgG2_Beta Spike
ADCP_Spike
IgA_S1
LV1 loadings
enriched in
2nd
3rd
r
1.0
0.5
0.0
−0.5
−1.0
ADCP_Spike
FcR2b_B.1.1.7.Spike
FcR2b_B.1.351.Spike
FcR3a_RBD
FcR2b_Spike
FcR3a_B.1.1.7.Spike
FcR3a_Delta.Spike
ADNP_Spike
FcR2a_B.1.1.7.Spike
FcR2b_P1.Spike
FcR2a_Spike
FcR3a_B.1.351.Spike
FcR3a_P1.Spike
FcR2a_B.1.351.Spike
FcR2a_P1.Spike
FcR3a_Spike
FcR2b_RBD
FcR2b_Delta.Spike
IgG2_Delta.Spike
IgG2_P1.Spike
IgG2_B.1.351.Spike
IgG2_B.1.1.7.Spike
IgG2_Spike
IgA_B.1.351.Spike
IgA_P1.Spike
IgA_B.1.1.7.Spike
IgA_Delta.Spike
IgA_Spike
BNT162b2
FcR2b_Alpha Spike
IgM_Alpha Spike
IgG3_RBD
CD107a_Spike
IgM_RBD
IgM_S2
IgM_S1
MIP1b_Spike
IFNg_Spike
IgM_Spike
IgM_B.1.1.7.Spike
IgM_P1.Spike
IgM_B.1.351.Spike
IgG3_Spike
IgG3_Delta.Spike
FcR2b_B.1.1.7.Spike

Figure S2. Vaccine-induced antibody titer and function in maternal blood by trimester of vaccination and features enriched by trimester within vaccine type.

(A) Spike-specific IgG2, IgG3, IgM and IgA were measured by Luminex. The dot plots show the titer for mothers who received vaccination in the 1$^{st}$ (orange), 2$^{nd}$ (blue), or 3$^{rd}$ (pink) trimester.

(B) The dot plots show the Spike-specific antibody-dependent NK cell activation, as measured by % MIP-1b + and % IFN-g+ NK cells, in maternal blood, based on trimester of vaccination. Significance was determined by Kruskal-Wallis test followed by posthoc Benjamini-Hochberg correction adjustment, * $p < 0.05$, ** $p < 0.01$, *** $p < 0.001$.

(C) The bar plot shows the LV1 of LASSO-selected features for the PLSDA built in Figure 2E. The color represents the trimester in which the feature was enriched. The network shows the features that are significantly correlated with LASSO-selected features ($|r| > 0.8$, $p < 0.05$). Lines connect significantly correlated features.

(D) The bar plot shows the LV1 of LASSO-selected features for the PLSDA built in Figure 2F. The color represents the trimester in which the feature was enriched. The network shows the features that are significantly correlated with LASSO-selected features ($|r| > 0.8$, $p < 0.05$). Lines connect significantly correlated features.

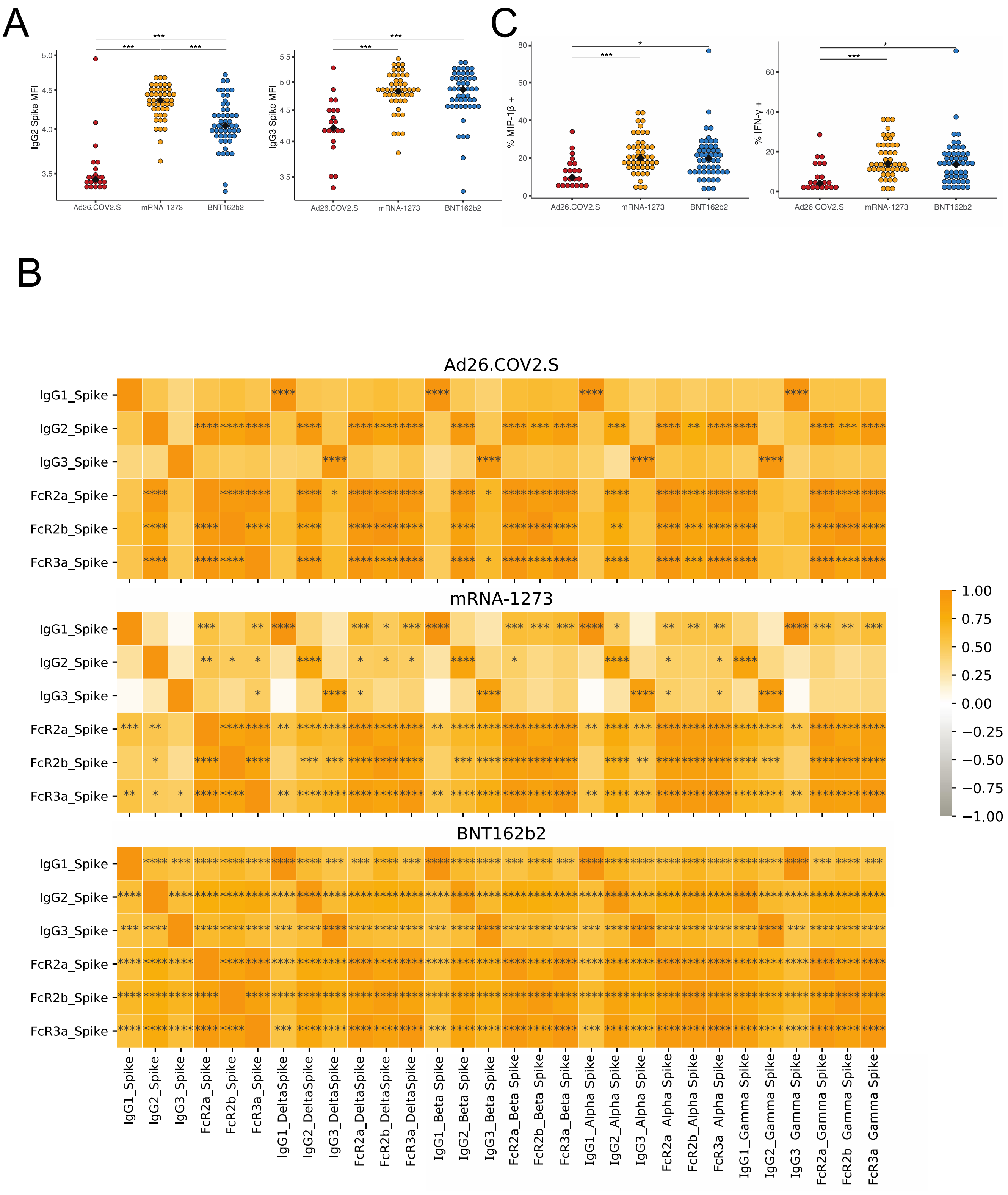
A
IgG2 Spike MFI
IgG3 Spike MFI
Ad26.COV2.S
mRNA-1273
BNT162b2
C
% MIP-1β +
% IFN-γ +
B
Ad26.COV2.S
mRNA-1273
BNT162b2
IgG1_Spike
IgG2_Spike
IgG3_Spike
FcR2a_Spike
FcR2b_Spike
FcR3a_Spike
IgG1_DeltaSpike
IgG2_DeltaSpike
IgG3_DeltaSpike
FcR2a_DeltaSpike
FcR2b_DeltaSpike
FcR3a_DeltaSpike
IgG1_Beta Spike
IgG2_Beta Spike
IgG3_Beta Spike
FcR2a_Beta Spike
FcR2b_Beta Spike
FcR3a_Beta Spike
IgG1_Alpha Spike
IgG2_Alpha Spike
IgG3_Alpha Spike
FcR2a_Alpha Spike
FcR2b_Alpha Spike
FcR3a_Alpha Spike
IgG1_Gamma Spike
IgG2_Gamma Spike
IgG3_Gamma Spike
FcR2a_Gamma Spike
FcR2b_Gamma Spike
FcR3a_Gamma Spike
1.00
0.75
0.50
0.25
0.00
−0.25
−0.50
−0.75
−1.00

Figure S3. Vaccine type results in different vaccine-induced antibody titer and function in cord blood.

(A) Spike-specific IgG2 and IgG3 were measured by Luminex. The dot plots show the titer for cords whose mothers received Ad26.COV2.S (red), mRNA-1273 (yellow) or BNT162b2 (blue).

(B) The heatmaps show the spearman correlation of antibody features against the Spike of the ancestral virus versus antibody features against the Spikes from variants of concern in maternal plasma, analyzed separately for each vaccine platform. Orange indicates a positive correlation, * $p < 0.05$, ** $p < 0.01$, *** $p < 0.001$, **** $p < 0.0001$

(C) The dot plots show the Spike-specific antibody-dependent NK cell activation, as measured by % MIP-1b + and % IFN-g+ NK cells, in cord blood. Significance was determined by Kruskal-Wallis test followed by posthoc Benjamini-Hochberg correction adjustment, * $p < 0.05$, ** $p < 0.01$, *** $p < 0.001$.

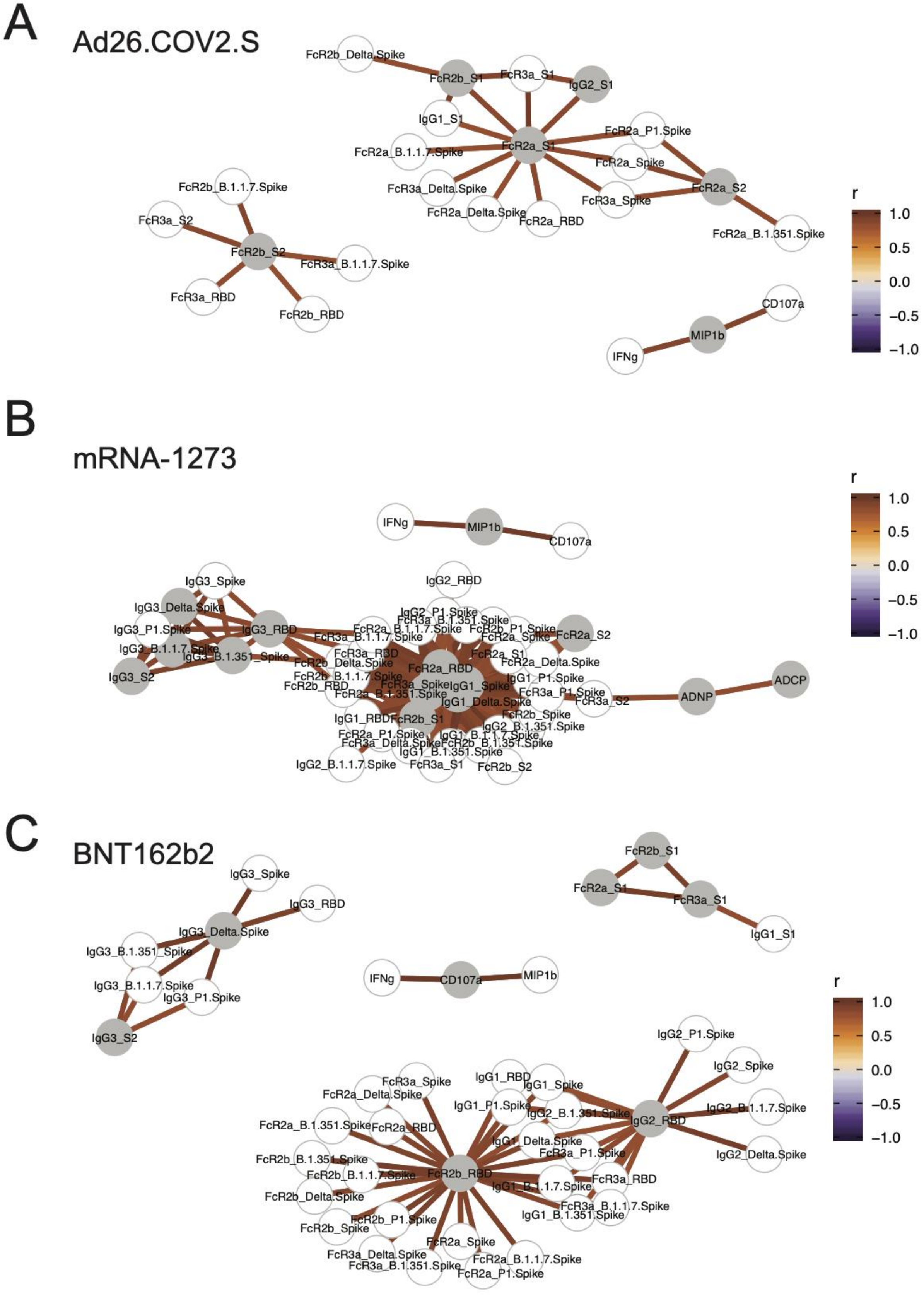
A
Ad26.COV2.S
FcR2b_Delta.Spike
FcR2b_S1
FcR3a_S1
IgG2_S1
IgG1_S1
FcR2a_P1.Spike
FcR2a_B.1.1.7.Spike
FcR2a_S1
FcR2a_Spike
FcR3a_Delta.Spike
FcR2a_S2
FcR3a_Spike
FcR2a_Delta.Spike
FcR2a_RBD
FcR2a_B.1.351.Spike
FcR2b_B.1.1.7.Spike
FcR3a_S2
FcR2b_S2
FcR3a_B.1.1.7.Spike
FcR3a_RBD
FcR2b_RBD
CD107a
MIP1b
IFNg
r
1.0
0.5
0.0
−0.5
−1.0
B
mRNA-1273
IFNg
MIP1b
CD107a
IgG2_RBD
IgG3_Spike
IgG3_Delta.Spike
IgG3_P1.Spike
IgG3_RBD
IgG3_B.1.1.7.Spike
IgG3_B.1.351.Spike
IgG3_S2
IgG2_P1.Spike
FcR3a_B.1.351.Spike
FcR2a_B.1.1.7.Spike
FcR2b_P1.Spike
FcR2a_S2
FcR3a_B.1.1.7.Spike
FcR2a_Spike
FcR3a_RBD
FcR2a_S1
FcR2b_Delta.Spike
FcR2a_RBD
FcR2a_Delta.Spike
FcR2b_B.1.1.7.Spike
IgG1_P1.Spike
FcR2b_RBD
FcR3a_Spike
IgG1_Spike
FcR2a_B.1.351.Spike
FcR3a_P1.Spike
IgG1_Delta.Spike
FcR3a_S2
ADNP
ADCP
IgG1_RBD
FcR2b_S1
FcR2b_Spike
IgG2_B.1.351.Spike
FcR2a_P1.Spike
IgG1_B.1.1.7.Spike
FcR3a_Delta.Spike
FcR2b_B.1.351.Spike
IgG1_B.1.351.Spike
IgG2_B.1.1.7.Spike
FcR3a_S1
FcR2b_S2
r
1.0
0.5
0.0
−0.5
−1.0
C
BNT162b2
IgG3_Spike
IgG3_RBD
IgG3_Delta.Spike
IgG3_B.1.351_Spike
IgG3_B.1.1.7.Spike
IgG3_P1.Spike
IgG3_S2
FcR2b_S1
FcR2a_S1
FcR3a_S1
IgG1_S1
IFNg
CD107a
MIP1b
r
1.0
0.5
0.0
−0.5
−1.0
IgG2_P1.Spike
IgG2_Spike
IgG2_B.1.1.7.Spike
IgG2_RBD
IgG2_Delta.Spike
FcR3a_Spike
IgG1_RBD
IgG1_Spike
FcR2a_Delta.Spike
IgG1_P1.Spike
IgG2_B.1.351.Spike
FcR2a_B.1.351.Spike
FcR2a_RBD
IgG1_Delta.Spike
FcR3a_P1.Spike
FcR2b_B.1.351.Spike
FcR2b_B.1.1.7.Spike
FcR2b_RBD
FcR3a_RBD
IgG1_B.1.1.7.Spike
FcR2b_Delta.Spike
FcR3a_B.1.1.7.Spike
IgG1_B.1.351.Spike
FcR2b_P1.Spike
FcR2b_Spike
FcR2a_Spike
FcR3a_Delta.Spike
FcR2a_B.1.1.7.Spike
FcR3a_B.1.351.Spike
FcR2a_P1.Spike

Figure S4. Vaccine-induced antibodies transferred to cord are highly coordinated.

(A-C) The network shows the antibody features that were significantly correlated ($|r| > 0.8$, $p < 0.05$) with the LASSO-selected features for the mPLSDA built in Figure 4C (A), Figure 4D (B), Figure 4E (C). Lines connect significantly correlated features.
